# Retrospective SARS-CoV-2 Real-Time PCR Testing of Stored Bronchoalveolar Lavage Samples from February 2020

**DOI:** 10.1101/2020.08.03.20167320

**Authors:** Douglas Challener, Aditya Shah, Matthew Binnicker, Andrew Badley, John O’Horo

## Abstract

Bronchoalveolar lavage samples (n=34) collected in February, 2020 prior to the wide availability of molecular testing for SARS-CoV-2 were retrospectively assayed for presence of viral RNA. None of these patients qualified for SARS-CoV-2 testing based on Centers for Disease Control criteria at the time. None of the samples tested positive for SARS-CoV-2, suggesting that the virus was not yet widespread in Minnesota at the time these samples were obtained.

## Introduction

As the global COVID-19 pandemic progresses, there is increasing interest in determining whether SARS-CoV-2 was circulating prior to the availability of widespread testing. Testing for COVID-19 in the United States was initially very limited, and early testing guidelines from the Centers for Disease Control and Prevention (CDC) recommended testing only patients with symptoms. However, it is now known that there are a significant proportion of people who are infected but remain asymptomatic.^1^ This has led to concern that early cases of COVID-19 may have been missed, and that the virus might have been circulating throughout the country prior to the first reported cases.

The timing of the arrival of SAR-CoV-2 in the United States remains unclear. The first case in the United States was detected in Washington State on January 20, 2020 in a 35-year-old who had recently travelled to visit family in Wuhan, China.^2^ Retrospective testing of influenza-swabs collected between January and March, 2020 from 3,524 participants in the Seattle, Washington region revealed a positive sample that had been obtained on February 24, 2020.^3^ Detection of community transmission of SARS-2-CoV in the United States at the end of February prompted the CDC to expand testing criteria, previously limited to only those with recent travel to Wuhan City, China, or close contact with a laboratory-proven case or patient under investigation (PUI).^4^ The first known death in the United States from COVID-19 was on February 6, 2020 in a patient with no known risk factors for exposure in Santa Clara County, California, USA. This suggests that there was community spread of the virus in Northern California in late January, 2020.^5,6^

This study assessed whether SARS-CoV-2 was circulating in the upper Midwest prior to the initiation of broad testing for the virus. To address this question, stored lower respiratory tract specimens collected at the Mayo Clinic in Rochester, MN in February, 2020 were identified and tested for SARS-CoV-2 RNA using an emergency use authorized (EUA) real-time PCR assay.

## Methods

Bronchoalveolar lavage (BAL) samples (n=34) that had been collected during the month of February, 2020 in Rochester, Minnesota and stored frozen at −20°C were tested by a real-time reverse transcriptase (RT)-PCR to detect SARS-CoV-2 RNA. Viral RNA was extracted from these samples using the bioMérieux easyMAG/eMAG, with subsequent PCR amplification on the Roche LightCycler 480. The assay targets two gene regions within SARS-CoV-2; the open reading frame (Orf1ab) and nucleocapsid protein (N) genes.^7^ This method has received emergency use authorization (EUA) from the Food and Drug Administration and was validated for testing of BAL samples.

A physician reviewed the electronic medical record to obtain the original indication for the BAL. The clinical indications for the procedure were categorized into acute (<4 weeks) and chronic symptoms (>4 weeks). We also determined if the patient met clinical criteria for suspected COVID-19 (based on the CDC surveillance case definition approved on April 5, 2020) at the time of their procedure as well as their final diagnosis. Descriptive statistics of the cohort were generated with JMP Pro 14 (SAS Institute Inc., Cary, N). This study was approved by the institutional review board at our center.

## Results

Stored BAL samples from 34 patients were evaluated for SARS-CoV-2 RNA. Of these patients, 23 (67%) of the patients were residents of Minnesota and 18 (53%) were male. The samples were collected between February 6, 2020 and February 20, 2020. Twenty-three (67%) samples were collected while the patient was admitted to the hospital and the rest were obtained in the outpatient setting. Twenty one (62%) of the samples were obtained to evaluate for an acute respiratory process, while 4 (12%) samples were from patients in whom infection was not being considered as part of the differential diagnosis. These were samples collected on an inpatient (n = 23) and outpatient (n = 11) basis for a variety of indications including respiratory failure (mainly in an inpatient setting) and evaluation of chronic lung imaging abnormalities (mainly in an outpatient setting). Six of the patients met clinical case definition criteria for suspected COVID-19. Eighteen patients were diagnosed with an infection (Table 1). All 34 samples tested negative for SARS-CoV-2 RNA via PCR.

**Table 1.** Final Clinical Diagnosis Following Bronchoalveolar Lavage

| **Diagnostic Category** | **Number (n=34)** |
| --- | --- |
| Infection |  |
| Fungal | 5 |
| Viral | 4 |
| Bacterial | 7 |
| Mycobacteria | 2 |
| Autoimmune | 4 |
| Hypersensitivity<br>Pneumonitis | 2 |
| Anatomical<br>abnormality | 1 |
| Malignancy | 1 |
| Other or Unknown | 6 |

## Discussion

In our study, there was no evidence of SARS-CoV-2 RNA in the 34 stored BAL samples that were obtained in February of 2020. This suggests that there was not widespread community circulation of SARS-CoV-2 in Rochester, MN prior to the availability of widespread testing. Multiple previous studies have evaluated stored nasopharyngeal swabs originally performed for evaluation of influenza; however, none have evaluated stored lower respiratory tract samples. Lower respiratory tract samples such as bronchoalveolar lavage may be more sensitive than samples from the upper respiratory tract.^8^ This cohort also represents patients who were more ill than previous prevalence studies with most being hospitalized at the time of their sample collection. Despite the limited sample size, the lack of any positive tests is useful as further work is performed to characterize the initial stages of the COVID pandemic in the United States.

We acknowledge Aimee Boerger who helped identify the samples used in this study.

## Data Availability

There is no additional data that is not included in the manuscript itself.

